# Diagnostic accuracy and usability of the Pluslife MTB assay on tongue and sputum swabs in symptomatic and asymptomatic adults in South Africa

**DOI:** 10.64898/2026.06.28.26350594

**Authors:** Anura David, Lesley Erica Scott, Lyndel Singh, Puleng Marokane, Manuel Pedro da Silva, Danielle Gast, Lara Noble, Ziyaad Waja, Tumelo Moloantoa, Neil Martinson, Wendy Stevens

## Abstract

**Background:** Access to accurate tuberculosis (TB) diagnostics remains limited, particularly in high-burden settings. The Pluslife MTB assay is among the first molecular tests specifically designed for swab-based detection of *Mycobacterium tuberculosis* complex (MTBC) and offers near point-of-care use.

**Methods:** We conducted a prospective diagnostic accuracy study in South Africa to evaluate the performance of the Pluslife assay on tongue swabs (TSs) and sputum swabs (SSs) among symptomatic and asymptomatic adults. Results were compared against liquid culture as the reference standard and Xpert MTB/RIF Ultra (Xpert Ultra) as a comparator. Operational characteristics and ease-of-use were assessed through structured observation and Likert-scale scoring by testing personnel.

**Results:** Of 256 participants enrolled, 92 [36%] were people with HIV (PHIV) and 217 were included in the final analysis. Culture confirmed TB in 41/217 (19%). The Pluslife assay demonstrated sensitivity of 85% (95% CI: 70.8-94.4) on SSs, comparable to Xpert Ultra on sputum (83%, 95% CI: 67.9-92.8), and detected one additional case missed by Xpert Ultra. Sensitivity on TSs was lower (63%, 95% CI: 46.9-77.9), particularly among PHIV. Specificity exceeded 97% across specimen types. Concordance on TSs between Pluslife and Xpert Ultra increased with higher bacterial loads. Operational evaluation showed short hands-on time and high ease-of-use scores, though limitations were noted for patient identifier recording and troubleshooting on the Pluslife MiniDock device.

**Conclusions:** The Pluslife MTB assay on SSs shows comparable performance to existing rapid diagnostics and favorable usability. Tongue swabs remain feasible but less reliable, supporting sputum as the preferred first specimen for TB diagnosis.

**Summary:** The Pluslife MTB assay showed high diagnostic accuracy on sputum swabs, comparable to Xpert Ultra, with high specificity and acceptable usability. Performance on tongue swabs was lower, particularly among PHIV, supporting sputum as the preferred first specimen for TB diagnosis.

## Background

Of the 8.3 million people who were reported as newly diagnosed with TB in 2024, only 54% were initially tested with a WHO-recommended rapid test (WRD) (1). Barriers to the uptake of available technologies include factors such as high costs, supply chain issues, lack of local technical support, requirements for equipment maintenance, human resource and training gaps and health system and infrastructure constraints (2, 3). Therefore, improving access to accurate diagnostics remains essential for strengthening TB control. The growing diagnostic pipeline, along with the WHO’s shift to endorsing classes of diagnostics, offers countries increased flexibility to tailor testing approaches to both centralised and decentralised service-delivery models (4, 5) for TB. The most recent definitions now distinguish between automated and manual low-complexity (LC) nucleic acid amplification tests (NAATs) (2). This shift is intended to enable TB testing closer to the patient at a reduced cost. The latest Tuberculosis Diagnostics Pipeline report (6) from the Treatment Action Group also highlights the emergence of near point-of-care (nPOC) technologies and the need for non-sputum specimens. Oral swabs, particularly tongue swabs, are a more accessible specimen that may improve case detection for individuals who are unable to produce sputum (7) and have been recommended by the WHO for TB diagnosis (8). Most swab evaluation studies thus far have paired downstream testing of swabs on existing technologies designed for sputum (9, 10) or on in-house polymerase chain reaction (PCR) assays (11).

The Pluslife MTB assay (Pluslife Biotech, Guangzhou, China) is among the first molecular tests specifically developed for swab-based *Mycobacterium tuberculosis* complex (MTBC) detection. Designed for nPOC use, it can analyze either tongue swabs (TSs) or sputum swabs (SSs). Preliminary studies, conducted primarily in settings with HIV prevalence of ~20% (12, 13), have reported sensitivities of 74–86% for TSs and 86–94% for SSs, with performance varying by symptom status and among people with HIV (PHIV) (14). However, comprehensive evaluations of the assay’s diagnostic performance, operational characteristics, and usability across diverse populations remain limited.

This study aimed to evaluate the diagnostic accuracy of the Pluslife MTB assay on TSs and SSs in both symptomatic and asymptomatic adults in Johannesburg, South Africa. Additionally, we assessed the operational characteristics and usability of the assay, including workflow efficiency, ease-of-use, and training requirements, to inform its potential integration into routine clinical and screening settings.

## Methods

### Study design

This cross-sectional, prospective diagnostic accuracy study evaluated the performance of the Pluslife MTB assay for detection of MTBC on TSs and SSs. Results were compared with a Mycobacterial Growth Indicator Tube (MGIT) liquid culture reference (Becton, Dickinson and Company, Sparks, MD, USA) and to Xpert MTB/RIF Ultra (Xpert Ultra; Cepheid, Sunnyvale, CA, USA) as a comparator, both performed on sputum.

Assay ease-of-use was assessed using a Likert scale completed by all testing personnel. A five-point scale was used to express level of agreement with each aspect evaluated: 1 (very difficult), 2 (difficult), 3 (neutral), 4 (easy), and 5 (very easy), with higher scores indicating more favourable ease-of-use characteristics. At least two operators completed the assessment, and an average score was assigned for each characteristic, followed by a category and overall ease-of-use score.

Operational characteristics of the Pluslife MTB assay and associated instrumentation were also evaluated. Parameters assessed included: hands-on time, procedural steps, ease of use, result interpretation, connectivity, training requirements, instrument performance, waste handling, maintenance needs, and shipping and storage conditions. Data were collected through direct observation of workflows, review of instrument functionality, and structured feedback from testing personnel.

### Ethics statement

Ethics approval for this study was obtained from the University of the Witwatersrand Human Research Ethics Committee (220612B). The trial was registered with the South African National Clinical Trials Registry (DOH-27-062023-6719). All participants provided written informed consent.

### Participant recruitment and study procedures

Adults were ambulatory, aged ≥18 years who were symptomatic or eligible for targeted universal testing for TB (TUTT) (15) and undergoing TB assessment were enrolled from 7 to 29 July 2025 at primary healthcare facilities: in Soweto and Matlosana Municipality, North West Province. The WHO-recommended four-symptom screen (cough, fever, weight loss, and night sweats) was administered to all participants. Participant characteristics including HIV status, TB history, diabetes mellitus status, medical conditions and demographic information were also collected. Two sputum specimens and two TSs were collected during a single study visit (Figure 1). The TSs were collected using the procedure described by Andama et al. (16), before sputum collection, with participants refraining from oral intake for at least 30 minutes prior. Briefly, TS collection from each participant was performed by research nurses by swabbing the dorsum of the tongue with a Copan FLOQSwab with 80mm breakpoint (Copan, Brescia, Italy) for 30 seconds, as far back as possible without initiating a gag reflex. The two sputum specimens were collected after three cycles of three forced cough efforts unless a mucoid specimen was produced after the first effort. Sputum specimens were randomly assigned to either routine testing on Xpert Ultra or research testing using SS and tested on the Pluslife MTB assay.

**Figure 1.**
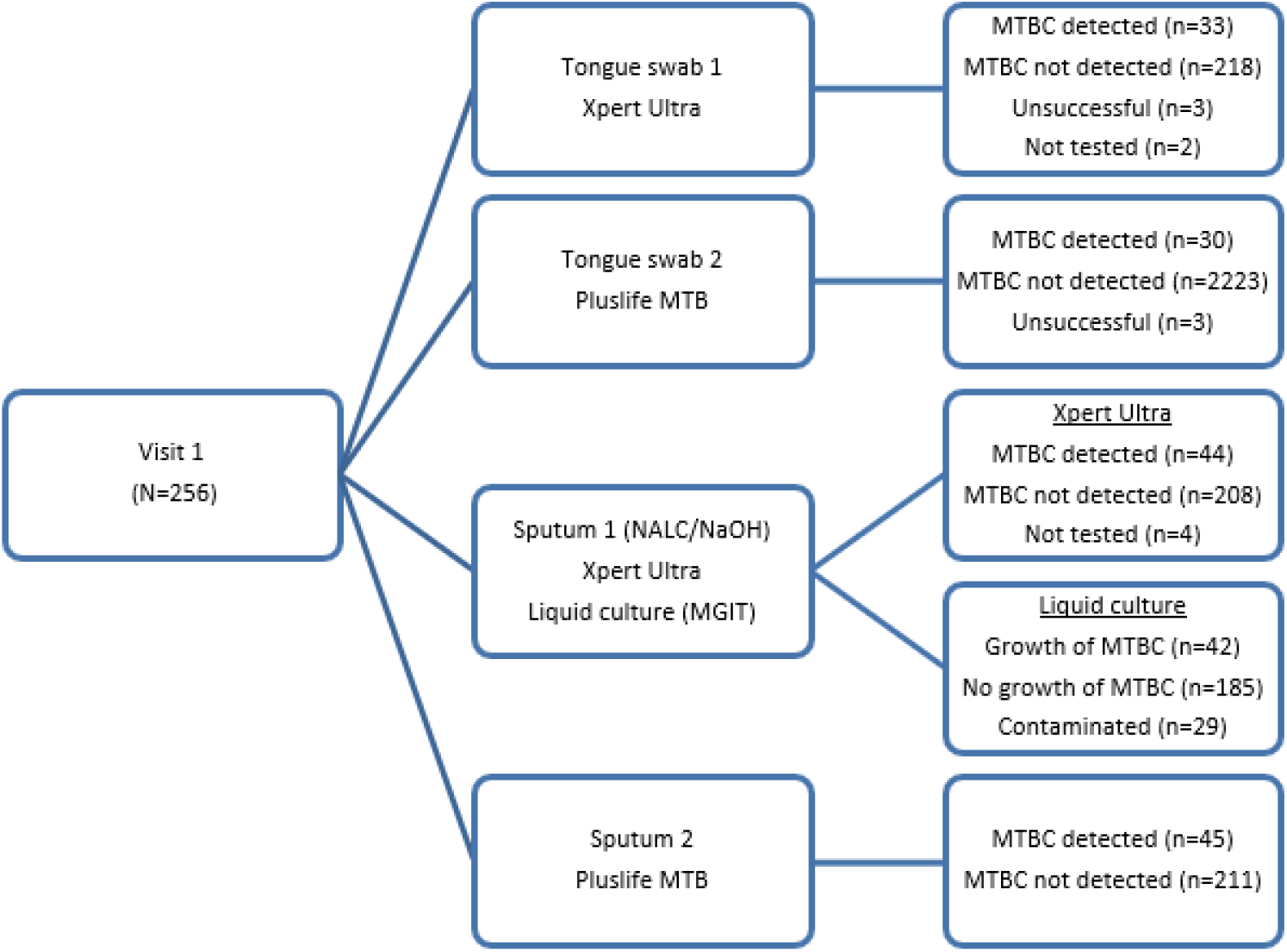
Specimen collection and diagnostic testing workflow at Visit 1. Participants provided two tongue swabs and two sputum specimens. Tongue swabs were tested using Xpert Ultra and Pluslife MTB assays, while sputum specimens underwent Xpert Ultra, liquid culture (Mycobacterial Growth Indicator Tube [MGIT]), and Pluslife MTB testing. MTBC, *Mycobacterium tuberculosis* complex

Specimens collected for routine and research testing were transported to the Wits Diagnostic Innovation Hub (WitsDIH) laboratory in Johannesburg.

### Routine testing

Routine testing and result return was performed by laboratory staff as per the current National Tuberculosis Management Guidelines (17). Firstly, sputum specimens were processed using N-acetyl-L-cysteine/Sodium hydroxide (NALC/NaOH) for MGIT liquid culture using adapted procedures from the MGIT^™^ procedure manual (18) and the BACTEC MGIT 960 System User’s Manual. The decontaminated sputum was then tested on Xpert Ultra according to manufacturer instructions while TS were processed using the diluted SR protocol by Ahls et al (19). Staff performing routine testing were blinded to results from the Pluslife MTB assay.

### Pluslife MTB testing

Sputum swabs and TSs were tested in the laboratory, by research personnel, as per manufacturer instructions, within 36 hours of collection. Specimens were stored at 2-8°C while awaiting testing. Testing personnel were blinded to routine testing results. For SS preparation in a biosafety cabinet, a clean swab (provided in the kit) was swirled 10 times in the sputum for ~10 seconds to allow the swab to be “dipped” in the sputum before processing. Residual sputum was decontaminated and tested as described above. For processing of both TSs and SSs, the swab was added to the tube containing the nucleic acid releasing agent, lysed for 5 minutes using the Thermolyse device (Pluslife), followed by testing on either the PM001 (MiniDock) or the PM008 analyser. The PM001 device can test a single specimen at a time while the PM008 format allows testing of eight specimens at a time. Results are interpreted as Positive, Negative or Invalid.

Any participant who tested positive on any assay was re-contacted. Those who were positive on liquid culture or Ultra were immediately referred, with a copy of their results, for initiation of TB treatment if it had not already begun. For participants who tested positive only on the Pluslife MTB assay, and who were not yet on TB treatment, a second confirmatory sputum specimen was obtained and sent to routine TB diagnostic services. If this confirmatory test was positive, the patient was referred for initiation of TB treatment.

### Outcomes and statistical analysis

For performance evaluation, SSs and TSs results from Pluslife MTB were compared with the gold standard liquid culture for MTBC detection. Data analysis included calculation of sensitivity, specificity, positive predictive value (PPV) and negative predictive value (NPV), with 95% confidence intervals (CI) calculated using the Wilson score method. Pluslife MTB results were additionally compared with Xpert Ultra on sputum.

The target sample size for the study was 200 participants, estimated using the standard sample size formula s: (n) = Z^2^ X p(1-p) / x^2^, where *Z* = 1.96, & where *p* = population proportion & where *x* = confidence interval as a proportion. Using these parameters, a sample size of 200 participants corresponds to a margin of error of approximately 6.9%, providing sufficient precision for estimating the primary outcomes of the study. To account for potential non-response or participant dropout, the sample size was increased by 10%, resulting in a final target enrolment of 220 participants ensuring adequate statistical power despite possible attrition during the study. To determine the performance of the Pluslife MTB assay using both TSs and SSs, only specimens that generated valid results across all tests (Pluslife MTB, Ultra and liquid culture) were included in the statistical analysis. Analyses were conducted in Stata version 16 (StataCorp, College Station, TX).

## Results

### Study population characteristics

Of 256 participants screened for enrolment over a 1-month period, the average age of participants was 38 years and 133/256 (52%) were male (Table 1). A total of 92/255 (36%) were PHIV with one participant reporting an unknown HIV status. Previous TB was reported by 39/256 (15%) participants. Most recruited participants were symptomatic (215/256 [84%]). Cough was the most reported symptom, experienced by 207/215 (96%) of participants with haemoptysis being the least reported on 12/215 (6%). A total of 3/256 (1%) participants showed elevated random glucose levels. Overall, 39/256 (15%) specimens had at least one unsuccessful test result (on Xpert Ultra, Pluslife MTB and/or culture). Of those with unsuccessful cultures, MTBC was detected in four participants using sputum and/or TS NAAT. Statistical performance analysis was conducted on test results from 217 participants.

**Table 1:** Demographic, clinical, and symptom characteristics of study participants.

| Characteristic | All (n=256) |
| --- | --- |
| *Demographics* |  |
| Age, mean (range), years | 38 (18-73) |
| Male sex, n (%) | 133 (51.9) |
| *HIV-related information* |  |
| People with HIV, n (%) | 92 (36.0) |
| HIV-negative, n (%) | 163 (63.9) |
| Unknown, n (%) | 1 (0.4) |
| Receiving <sup>a</sup>ARV therapy, n/total (%) | 84/88 (95.5) |
| Elevated Random glucose levels, n (%) | 3 (1.1) |
| *Co-morbidities* |  |
| Hypertension, n (%) | 19 (7.4) |
| Diabetes, n (%) | 8 (3.1) |
| *TB History* |  |
| Previously diagnosed with TB, n (%) | 39 (15.2) |
| *WHO four-symptom screen at presentation* |  |
| Cough, n/total (%) | 207/215 (96.2) |
| Nights sweats, n/total (%) | 89/215 (41.4) |
| Fever, n/total (%) | 80/215 (37.2) |
| Unexplained weight loss, n/total (%) | 52/215 (24.2) |
| *Other reported symptoms, n/total (%)* |  |
| Fatigue, n/total (%) | 60/215 (28.0) |
| Loss of appetite, n/total (%) | 55/215 (25.6) |
| Body pain, n/total (%) | 10/215 (4.7) |
| Haemoptysis, n/total (%) | 12/215 (5.6) |

<sup>a</sup>antiretroviral

### Diagnostic accuracy of the Pluslife MTB assay

Per bacteriological classification using culture, 41/217 (19%) were diagnosed with active TB disease (Table 2). The Pluslife MTB assay on SSs demonstrated better performance (85%) than on TSs (63%). Assay performance on SSs was similar to Xpert Ultra performance on sputum (83%) with Pluslife MTB identifying MTBC in one additional specimen (McNemar’s *p* = 1.000). On TS, Xpert Ultra was able to detect MTBC in three additional specimens compared to Pluslife MTB.

**Table 2:** The performance of Ultra and Pluslife MTB assays on tongue and sputum swabs compared to liquid culture for MTBC detection.

| Performance characteristic | Ultra result |  |  |  | Pluslife MTB result |  |  |  |
| --- | --- | --- | --- | --- | --- | --- | --- | --- |
|  | Sputum | n/N | TS | n/N | SS | n/N | TS | n/N |
| All (n=217) |  |  |  |  |  |  |  |  |
| Sensitivity % (95% CI) | 82.9 (67.9-92.8) | 34/41 | 70.7 (54.5-83.9) | 29/41 | 85.4 (70.8-94.4) | 35/41 | 63.4 (46.9-77.9) | 26/41 |
| Specificity % (95% CI) | 96.6 (92.7-98.7) | 170/176 | 99.4 (96.9-100) | 175/176 | 97.2 (93.5-99.1) | 171/176 | 98.9 (96.0-99.0) | 174/176 |
| PPV % (95% CI) | 85.0 (70.2-94.3) | 34/40 | 96.7 (82.8-99.9) | 29/30 | 87.5 (73.2-95.8) | 35/40 | 92.9 (76.5-99.1) | 26/28 |
| NPV % (95% CI) | 96.0 (92.0-98.4) | 170/177 | 93.6 (89.1-96.6) | 175/187 | 96.6 (92.8-98.7) | 171/177 | 92.1 (87.2-95.5) | 174/189 |
| Symptomatic participants only (n=166) |  |  |  |  |  |  |  |  |
| Sensitivity % (95% CI) | 87.5 (71.0-96.5) | 28/32 | 75.0 (56.6-88.5) | 24/32 | 90.6 (75.0-98.0) | 29/32 | 68.8 (50.0-83.9) | 22/32 |
| Specificity % (95% CI) | 97.0 (92.5-99.2) | 130/134 | 100 (97.3-100) | 134/134 | 97.8 (93.6-99.5) | 131/134 | 98.5 (94.7-99.8) | 132/134 |
| PPV % (95% CI) | 87.5 (71.0-96.5) | 28/32 | 100 (85.8-100) | 24/24 | 90.6 (75.0-98.8) | 29/32 | 91.7 (73.0-99.0) | 22/24 |
| NPV % (95% CI) | 97.0 (92.5-99.2) | 130/134 | 94.3 (89.1-99.5) | 134/142 | 97.8 (93.5-99.5) | 131/134 | 93.0 (87.3-96.5) | 132/142 |
| PHIV, irrespective of symptoms (n=67) |  |  |  |  |  |  |  |  |
| Sensitivity % (95% CI) | 75.0 (50.9-91.3) | 15/20 | 55.0 (31.5-76.9) | 11/20 | 80.0 (56.3-94.3) | 16/20 | 40.0 (19.1-63.9) | 8/20 |
| Specificity % (95% CI) | 93.6 (82.5-98.7) | 44/47 | 100 (92.5-100) | 47/47 | 95.7 (85.5-99.5) | 45/47 | 97.9 (88.7-99.9) | 46/47 |
| PPV % (95% CI) | 83.3 (77.8-96.6) | 15/18 | 100 (71.5-100) | 11/11 | 88.9 (65.3-98.6) | 16/18 | 88.9 (51.8-99.7) | 8/9 |
| NPV % (95% CI) | 89.8 (77.8-96.6) | 44/49 | 83.9 (71.7-92.4) | 47/56 | 91.8 (80.4-97.7) | 45/49 | 79.3 (66.6-88.8) | 46/58 |
| HIV-negative participants, irrespective of symptoms (n=127) |  |  |  |  |  |  |  |  |
| Sensitivity % (95% CI) | 88.2 (63.6-98.5) | 15/17 | 82.4 (56.6-96.2) | 14/17 | 88.2 (63.6-98.5) | 15/17 | 88.2 (63.6-98.5) | 15/17 |
| Specificity % (95% CI) | 99.1 (95.0-100) | 109/110 | 99.1 (95.5-100) | 109/110 | 98.2 (93.6-99.8) | 108/110 | 99.1 (95.0-100) | 109/110 |
| PPV % (95% CI) | 93.8 (69.8-99.8) | 15/16 | 93.3 (68.1-99.8) | 14/15 | 88.2 (63.6-98.5) | 15/17 | 93.8 (69.8-99.8) | 15/16 |
| NPV % (95% CI) | 98.2 (93.6-99.8) | 109/111 | 97.3 (92.4-99.4) | 109/112 | 98.2 (93.6-99.8) | 108/110 | 98.2 (93.6-99.8) | 109/111 |

TS, tongue swab; SS, sputum swab; n/N, the number of concordant results (n) out of the total number
of specimens in that category (N); CI, confidence intervals; PHIV, people with HIV

Table 3 summarizes the level of agreement between the Pluslife MTB assays on TSs and SSs and the Xpert Ultra semiquantitative categories, showing how concordance varied across different bacterial load levels.

**Table 3:** Comparison of Xpert Ultra semiquantitative results with Xpert Ultra tongue swab, Pluslife MTB TS and sputum swab.

| Xpert Ultra semiquantitative result (sputum) | Xpert Ultra TS, n (% agreement) | Pluslife MTB TS result, n (% agreement) | Pluslife MTB SS result, n (% agreement) |
| --- | --- | --- | --- |
| High (n=13) | 12 (92) | 13 (100) | 13 (100) |
| Medium (n=7) | 7 (100) | 6 (85.7) | 7 (100) |
| Low (n=13) | 10 (77) | 7 (53.8) | 13 (100) |
| Very low (n=1) | 0 (0) | 0 (0) | 1 (100) |
| Trace (n=6) | 0 (0) | 0 (0) | 1 (17) |

TS, tongue swab; SS, sputum swab; MTB, *Mycobacterium tuberculosis*; n, the number of concordant results for a particular test

### Unsuccessful test outcomes for Xpert Ultra and Pluslife MTB

The Xpert Ultra and Pluslife MTB assays each produced errors or invalid results in 3/256 (1.2%) TS tests, respectively. No unsuccessful results were observed on sputum with either assay. Although sufficient specimen volumes were available to allow repeat testing with the Pluslife MTB assay, these were not performed.

Overall, testing personnel reported that the Pluslife MTB system was straightforward to use across all operational categories (Table 4).

**Table 4:**
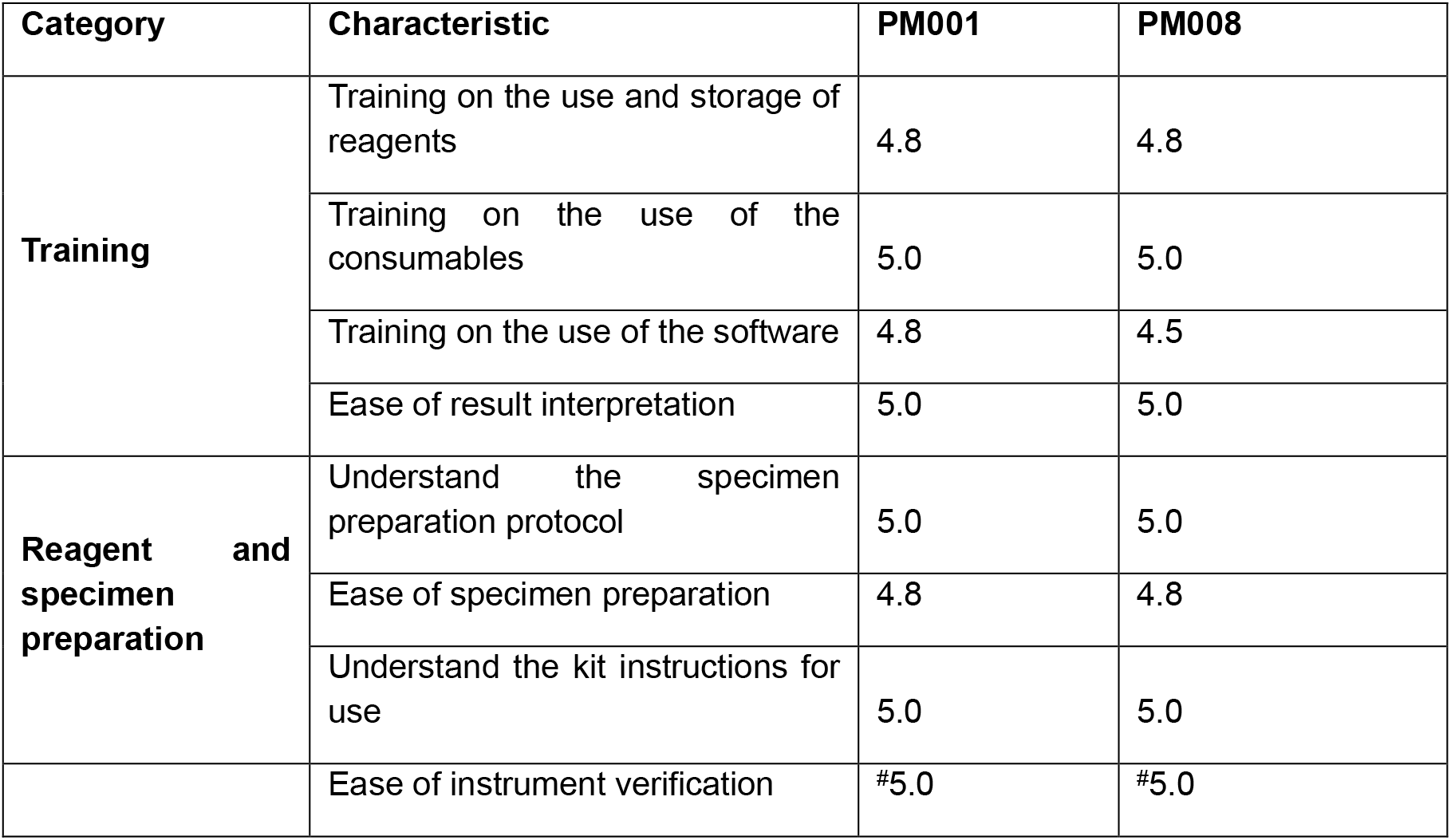
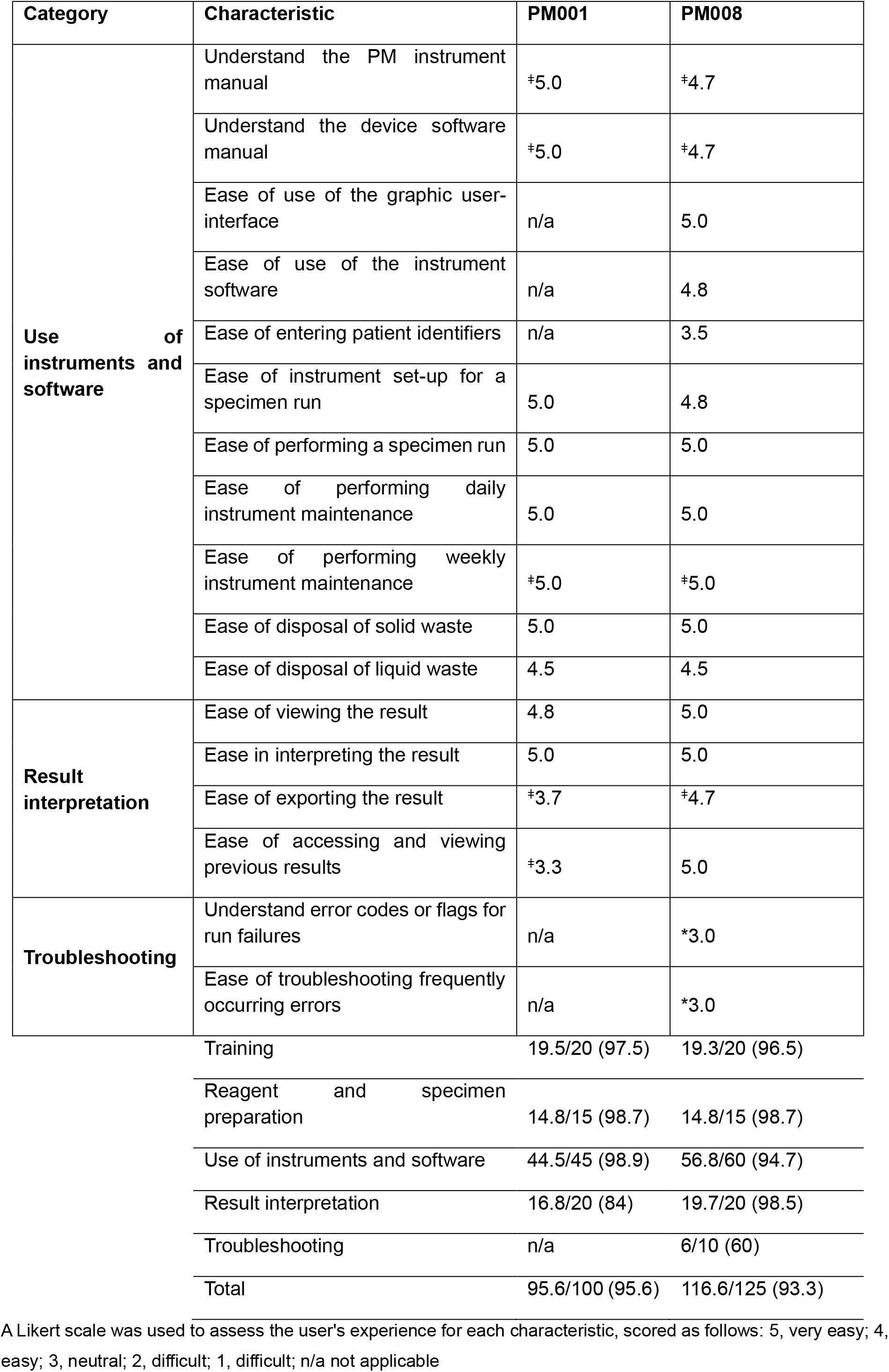
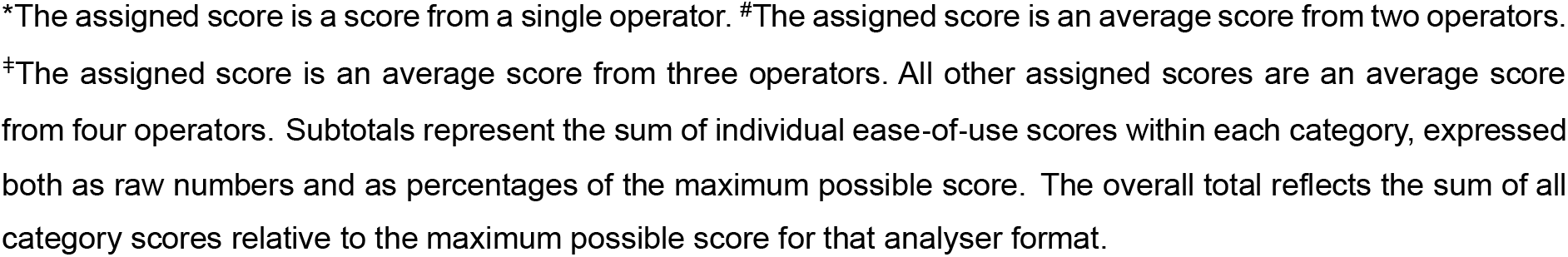
Ease-of-use table as scored by testing personnel for both analyser formats.

Operational considerations of the Pluslife MTB assay and instrumentation are summarised in Table 5.

**Table 5:** Operational considerations of the Pluslife MTB assay and instrumentation.

| Consideration | Comment |
| --- | --- |
| **Total Hands-on Time and Number of Steps** | Specimen processing for the MTB assay involves four steps: addition to sample release buffer (20 seconds), thermal lysis (5 minutes), addition of lysed material to testing cartridge (30 seconds) and testing on PM001/PM008 (12-25 minutes). Specimen registration on the PM008 is a manual process, hence the overall hands-on time was 2-3 minutes. |
| **Instruments** | Both the thermolyze instrument and PM001/PM008 devices have user-friendly instructions<br>The PM001 lacks a user interface, and patient identifiers cannot be entered. The user must manually record patient identifiers, cartridge serial numbers and other relevant information. |
| **Result interpretation** | At the end of the test run<br>the PM008 screen displays the test result with no interpretation required by the user<br>the PM001 uses a LED indicator to display the test result with no interpretation required by the user. The LED indicator light switches off once the test card is removed from the device so the user must record the result, prior to this. The instrument can be connected to a laptop, tablet or PC but since patient identifiers are not captured, the user needs to link the cartridge serial number and result to the patient. |
| **Connectivity** | The PM008 has an onboard printer and results can be printed. |
|  | The PM001 needs to be connected to a laptop, tablet or PC to view the results. Since patient identifiers cannot be captured on the device, these need to be manually recorded.<br>Wi-Fi connectivity can be used on the PM008 device to share results |
| **Training Needs** | Minimal technical training required; proficiency was achieved after 2-3 runs for a medical technologist and junior medical scientist. |
| **Instrument Failure Rate** | There was one occasion where a power outage caused failed runs.<br>Two modules on the PM008 device demonstrated invalid and false negative results which resolved on repeat on other modules. These modules were not used for the duration of the study, leaving 6/8 modules available for testing. |
| **Waste disposal** | The sample release tubes, residual sputum and swab shafts were discarded into biohazard waste boxes for incineration prior to disposal.<br>Test cards were discarded into sharps containers.<br>The foil seals, swab pouches and package inserts were discarded into normal waste<br>The kit cardboard boxes were sent for recycling |
| **Maintenance and Customer Support Needs** | Both the thermolyze device and the PM00 devices require minimal maintenance (cleaning the external surfaces of the instruments and discarding used cartridges). Customer support is easily accessible and responsive. |
| **Shipping and Storage** | The Pluslife MTB kits are transported and stored at room temperature. |

Operational considerations for implementation of the Pluslife MTB assay, assessed by up to four operators during the study. The table summarizes hands-on time, instrument usability, result interpretation, connectivity, training requirements, instrument reliability, waste disposal, maintenance, and shipping/storage needs.

## Conclusion

This study evaluated the performance of the Pluslife MTB assay using both TSs and SSs in a high TB burden setting. Participants were largely symptomatic, and a substantial proportion were living with HIV (36%), reflecting a population in which rapid diagnostic tools are most urgently needed. The assay demonstrated good diagnostic accuracy on SSs, performing similarly to Xpert Ultra and detecting one additional TB case. Other studies which evaluated TS performance on the Pluslife MTB assay showed an overall sensitivity of ≥76% (12-14) though up to 60% of participants in these populations had high bacterial burdens, as indicated by their Xpert Ultra semiquantitative results. These studies along with evaluations of other molecular assays (10, 20), consistently show that sensitivity on TS decrease, in specimens with low bacterial burden, when compared to sputum. In our study, where 50% of participants demonstrated low bacterial burden, overall sensitivity on TS was 63%, when compared to liquid culture - lower than previously reported (12-14). Another notable difference was the higher proportion of PHIV (36% versus ~20% in other studies), which may have contributed to the reduced TS performance. These findings suggest that TS are feasible but may be less reliable for routine diagnostic use, especially in paucibacillary disease.

When stratified by HIV status, diagnostic accuracy was reduced among PHIV, consistent with previous findings (21). Performance on SSs remained comparatively robust, while sensitivity on TS declined. These observations highlight the value of sputum-based testing even in populations where sputum collection can be challenging.

Concordance between Pluslife MTB and Xpert Ultra increased with higher bacterial loads, with 100% agreement observed for specimens reported as “high” or “medium” by Ultra. Reduced agreement at “low” and “trace” is expected for molecular assays and again mirrors the likelihood of fewer DNA targets at these concentrations. Tongue swabs were disproportionately affected by this trend, suggesting that sampling modality rather than assay chemistry may be responsible for the reduced sensitivity in low-burden disease.

The Pluslife MTB assay showed a low (1.2%) rate of unsuccessful tests and sufficient volume of lysed specimen is available for repeat testing. Training needs were minimal, with operators quickly gaining proficiency in specimen preparation, instrument operation, and result interpretation. Both the PM001 and PM008 devices were straightforward to install and use, and routine maintenance tasks were easily completed. Although result viewing and interpretation were generally intuitive, some limitations were identified for the PM001, particularly around recording identifiers and accessing or exporting results, and occasional troubleshooting difficulties were reported. Despite these platform-specific issues, overall usability scores reflected high operator satisfaction.

Hands-on time was short, with a simple four-step workflow for specimen processing. Both instruments were easy to operate, required limited training, and provided automated result read-outs. The absence of device interfaces meant that patient identifiers needed to be recorded manually for the PM001 device; however, connectivity options were available on the PM008. Waste management, routine maintenance, and storage needs were minimal, supporting feasibility for routine implementation in low-resource settings.

Overall, this laboratory evaluation supports the use of the Pluslife MTB assay on SSs as a viable alternative to existing rapid diagnostics. One disadvantage of the Pluslife MTB assay is that it does not provide resistance testing, which means that diagnostic algorithms need to be developed to address this gap. Implementation in decentralized settings will require attention to biosafety concerns, including the risk of sputum spills, aerosol release during SS preparation, and safe disposal of residual sputum following testing. While TSs may increase bacterial yield (22) and offer operational advantages, our findings confirm that sputum collection remains essential wherever possible to ensure diagnostic accuracy. Performance on TSs underscores the need for continued refinement of sampling strategies if this minimally invasive approach is to be widely adopted.

Given its operational simplicity, short hands-on time, and favorable usability scores, the Pluslife MTB assay may be particularly advantageous in resource-limited settings where laboratory infrastructure and skilled operators are scarce. Future work should focus on addressing the absence of resistance testing, while also informing biosafety practices and guiding workflow adaptations for safe and effective implementation.

## Data Availability

The datasets generated during this study cannot be made publicly available due to participant confidentiality. De-identified data may be made available from the corresponding author upon reasonable request, subject to approval by the relevant ethics committee.

## Acknowledgements

The authors thank the study participants; the Johannesburg and North West Health District for their support and collaboration on this study; and Guangzhou Pluslife Biotech Co., Ltd. (Pluslife) for providing technical support, kits, and associated instrumentation for this study. Pluslife was not involved in the study design and analysis and interpretation of results.

Conceptualization: A.D. and L.E.S.; data curation: Z.W., T.M. and N.M.; formal analysis: A.D.; funding acquisition: L.E.S. and W.S.; investigation: A.D., L.S., L.N., D.G., T.M., Z.W and P.M.; methodology: A.D. and L.E.S.; project administration: L.E.S., N.M. and W.S.; supervision: L.E.S.; validation: A.D.; visualization: A.D. N.M. and L.E.S.; writing (original draft): A.D.; writing (review and editing): A.D., L.E.S., P.M., M.P.D.S., L.S., D.G., L.N., Z.W., T.M., N.M. and W.S. All authors have read and agreed on the published version of the manuscript.

## References

1. Global Tuberculosis Report 2025. World Health Organisation; 2025[date accessed 28 November 2025]. Available from https://www.who.int/teams/global-programme-on-tuberculosis-and-lung-health/tb-reports/global-tuberculosis-report-2025.

2. Module 3: Diagnostics, Rapid Diagnostic for Tuberculosis Detection, WHO consolidated guidelines on Tuberculosis, World Health Organization; 2025[date accessed 02 May 2025]. Available from: https://www.who.int/publications/i/item/9789240107984.

3. Ntinginya NE, Kuchaka D, Orina F, Mwebaza I, Liyoyo A, Miheso B, Aturinde A, Njeleka F, Kiula K, Msoka EF, Meme H, Sanga E, Mwanyonga S, Olomi W, Minja L, Joloba M, Mmbaga BT, Amukoye E, Gillespie SH, Sabiiti W. Unlocking the health system barriers to maximise the uptake and utilisation of molecular diagnostics in low-income and middle-income country setting. BMJ Glob Health. 2021;6(8).

4. Module 2: Systematic screening for tuberculosis disease, WHO consolidated guidelines on Tuberculosis, World Health Organization; 2021 July[date accessed 22 November 2022]. Available from: https://www.who.int/publications/i/item/9789240022676

5. WHO. Target product profiles for tuberculosis diagnosis and detection of drug resistance; 2024[date accessed: 02 October 2024]. Available from: https://www.who.int/publications/i/item/9789240097698.

6. Pipeline Report 2025. Treatment Action Group. 2025.[date accessed: 28 November 2025]. Available from: https://www.eatg.org/hiv-news/tags-2025-pipeline-report-tb-diagnostics/.

7. Church EC, Steingart KR, Cangelosi GA, Ruhwald M, Kohli M, Shapiro AE. Oral swabs with a rapid molecular diagnostic test for pulmonary tuberculosis in adults and children: a systematic review. Lancet Glob Health. 2024;12:e45–54.

8. World Health Organization. WHO issues new recommendations on near point-of-care tests for TB diagnosis, including tongue swabs. Geneva: WHO; 2026.[date accessed: 27 February 2026]. Available from: https://www.who.int/teams/global-programme-on-tuberculosis-and-lung-health/diagnosis-treatment/npoc-tongue-swabs-and-sputum-pooling-for-tb

9. Wood RC, Luabeya AK, Dragovich RB, Olson AM, Lochner KA, Weigel KM, Codsi R, Mulenga H, de Vos M, Kohli M, Penn-Nicholson A, Hatherill M, Cangelosi GA. Tongue swab testing on two automated tuberculosis diagnostic platforms, Cepheid Xpert® MTB/RIF Ultra and Molbio Truenat® MTB Ultima. J Clin Microbiol. 2024;10(4).

10. David A, Singh L, da Silva MP, Peloakgosi-Shikwambani K, Nsingwane Z, Molepo V, Stevens W, Scott LE. Diagnostic Accuracy of the Cobas® MTB and Cobas MTB/RIF-INH Assays on Sputum and the Cobas MTB Assay on Tongue Swabs for Mycobacterium tuberculosis Complex Detection in Symptomatic Adults in South Africa. Biomedicines. 2025;13(10).

11. LaCourse SM, Seko E, Wood R, Bundi W, Ouma GS, Agaya J, Richardson BA, John-Stewart G, Wandiga S, Cangelosi GA. Diagnostic performance of oral swabs for non-sputum based TB diagnosis in a TB/HIV endemic setting. PLoS ONE. 2022;17(1):e0262123.

12. Mbuli C, Fokou TJB, Nsamenang R, Nestor B, Nguimfack G, Ndze MN, Konso J, Mana ZA, Arthur N, Nzebele TM, Denis N, Ndi N N, Wandji IAG, Fundoh M, Toussaint MG, Bello O, Paul M, Adeline F, Comfort V, Teyim P, Donfack VFD, Garg T, Creswell J, Sander M, Consortium RT. Diagnostic Performance of the Pluslife MiniDock MTB and Molbio MTB Ultima Assays to Detect Tuberculosis From Tongue and Sputum Swabs Among Outpatients and in Active Case Finding in Cameroon. Clin Infect Dis. 2025;ciaf709.

13. Steadman A, Kumar KM, Asege L, Kato-Maeda M, Mukwatamundu J, Shah K, Trang T, Ball A, Khimani K, Kim DDT, Michael JS, Christopher DJ, Phan H, Yerlikaya S, Nahid P, Denkinger CM, Cattamanchi A, Andama A. Diagnostic accuracy of swab-based molecular tests for tuberculosis using near-point-of-care platforms: a multi-country evaluation. EBioMedicine. 2025;121:105991.

14. Wu Z, Yan L, Lai X, Yang J, Liang J, Ma X, Xie J, Ji L, Wang Y, Hu J, Chen R, Lv C, Fu X, Chen Y, Guo H, Xu L, Liang H, Liu S, Chen Z, Huang M, Zhong X, Chen X. Diagnostic accuracy of a novel point-of-care tongue swab assay for pulmonary tuberculosis: a multicentre prospective study. Clin Microbiol Infect. 2025.

15. Martinson NA, Nonyane BAS, Genade LP, Berhanu RH, Naidoo P, Brey Z, Kinghorn A, Nyathi S, Young K, Hausler H, Connell L, Lutchminarain K, Swe Swe-Han K, Vreede H, Said M, von Knorring N, Moulton LH, Lebina L, team TT. Evaluating systematic targeted universal testing for tuberculosis in primary care clinics of South Africa: A cluster-randomized trial (The TUTT Trial). PLoS Med. 2023;20(5).

16. Andama A, Steadman AE, Ahls C, Cangelosi GA, David A, de Vos M, Heichman K, Kato-Maeda M, Penn-Nicholson A, Olson A, Scott L, Turnbull L, Wood R, Weigel K, Cattamanchi A. Consensus standard operating procedure for collection of tongue swabs for TB diagnostics 2024 [Available from: https://www.protocols.io/view/consensus-standard-operating-procedure-for-collect-kxygxyw54l8j/v1.

17. South African National Department of Health. National guideline on the treatment of tuberculosis infection. [date accessed: 21 August 2024]. [press release]. 2023.

18. FIND. MGIT Manual: Manual for BACTEC MGIT 960 TB System. Geneva: Foundation for Innovative New Diagnostics; 2006.[date accessed: 21 November 2022]. Available from: https://www.finddx.org/wp-content/uploads/2023/02/20061101_rep_mgit_manual_FV_EN.pdf

19. Ahls C, David A, Chilambi GS, Cattamanchi A, de Vos M, Heard K, Heichman K, Penn-Nicholson A, Scott L, Steadman A, Turnbull L, Alland D. Xpert MTB/RIF Ultra testing from tongue swabs-Diluted SR method. 2024 [Available from: https://www.protocols.io/view/xpert-mtb-rif-ultra-testing-from-tongue-swabs-dilu-14egn69nyl5d/v1.

20. Zhang F, Wang Y, Zhang X, Liu K, Shang Y, Wang W, Liu Y, Li L, Pang Y. Diagnostic accuracy of oral swab for detection of pulmonary tuberculosis: a systematic review and meta-analysis. Front Med (Lausanne). 2024;10.

21. David A, Singh L, Peloakgosi-Shikwambani K, Nsingwane Z, Molepo V, Cangelosi G, da Silva P, Stevens W, Scott L. Diagnostic accuracy of self-collected tongue swabs for Mycobacterium tuberculosis complex detection using Xpert MTB/RIF Ultra. Clin Microbiol Infect. 2025;31(6):1040–5.

22. Moe C A., Luswata RK, Barrameda AJ, L. H, Muzazu S, Crowder R, Andama AO, Denkinger CM, Muyoyeta M, Phan H, Cattamanchi A, Yu C. Diagnostic Yield of Tongue Swab-Compared to Sputum-Based Molecular Testing for Tuberculosis in Four High-Burden Countries. Clin Infect Dis. 2026; 82(4):e816–e823.

